# An open label, adaptive, phase 1 trial of high-dose oral nitazoxanide in healthy volunteers: an antiviral candidate for SARS-CoV-2

**DOI:** 10.1101/2021.09.10.21263376

**Authors:** Lauren E Walker, Richard FitzGerald, Geoffrey Saunders, Rebecca Lyon, Michael Fisher, Karen Martin, Izabela Eberhart, Christie Woods, Sean Ewings, Colin Hale, Rajith KR Rajoli, Laura Else, Sujan Dilly-Penchala, Alieu Amara, David G Lalloo, Michael Jacobs, Henry Pertinez, Parys Hatchard, Robert Waugh, Megan Lawrence, Lucy Johnson, Keira Fines, Helen Reynolds, Timothy Rowland, Rebecca Crook, Kelly Byrne, Pavel Mozgunov, Thomas Jaki, Saye Khoo, Andrew Owen, Gareth Griffiths, Thomas E Fletcher, on behalf of the AGILE platform

**Author notes:** **Author for correspondence:** Dr Lauren E Walker, Institute of Systems, Molecular and Integrative Biology (ISMIB), University of Liverpool, Block A: Waterhouse Building, 1-5 Brownlow Street, Liverpool L69 3GL, 0151 794 5407. **Conflicts of interest statement** AO is Director of Tandem Nano Ltd. AO has received research funding from ViiV, Merck, Janssen and consultancy from Gilead, ViiV and Merck not related to the current paper. Ridgeback and GlaxoSmithKline have provided funding to the AGILE phase I/II platform to evaluate SARS-CoV-2 candidates independently of the current trial. GG has received funding from Janssen-Cilag, Astra Zeneca, Novartis, Astex, Roche, Heartflow, Celldex, BMS, BionTech, Cancer Research UK, NIHR, British Lung Foundation, Unitaid, GSK for unrelated academic clinical trials and programme funding. SK has received funding from Merck, ViiV, Janssen, Gilead for unrelated academic trials. No other conflicts are declared by the authors. **Funding** This trial was funded by Unitaid as part of a supplement to project LONGEVITY in response to the SARS-CoV-2 pandemic. AO also acknowledges research funding from EPSRC (EP/R024804/1; EP/S012265/1), NIH (R01AI134091; R24AI118397), European Commission (761104). The authors also acknowledge funding from Wellcome Trust [221590/Z/20/Z] and Medical Research Council [MR/V028391/1] for the AGILE phase I/II platform trial.

## Abstract

Repurposing approved drugs may rapidly establish effective interventions during a public health crisis. This has yielded immunomodulatory treatments for severe COVID-19, but repurposed antivirals have not been successful to date because of redundancy of the target in vivo or suboptimal exposures at studied doses. Nitazoxanide is an FDA approved antiparasitic medicine, that physiologically-based pharmacokinetic (PBPK) modelling has indicated may provide antiviral concentrations across the dosing interval, when repurposed at higher than approved doses. Within the AGILE trial platform (NCT04746183) an open label, adaptive, phase 1 trial in healthy adult participants was undertaken with high dose nitazoxanide. Participants received 1500mg nitazoxanide orally twice-daily with food for 7 days. Primary outcomes were safety, tolerability, optimum dose and schedule. Intensive pharmacokinetic sampling was undertaken day 1 and 5 with Cmin sampling on day 3 and 7. Fourteen healthy participants were enrolled between 18^th^ February and 11^th^ May 2021. All 14 doses were completed by 10/14 participants. Nitazoxanide was safe and well tolerated with no significant adverse events. Moderate gastrointestinal disturbance (loose stools) occurred in 8 participants (57.1%), with urine and sclera discolouration in 12 (85.7%) and 9 (64.3%) participants, respectively, without clinically significant bilirubin elevation. This was self-limiting and resolved upon drug discontinuation. PBPK predictions were confirmed on day 1 but with underprediction at day 5. Median Cmin was above the in vitro target concentration on first dose and maintained throughout. Nitazoxanide administered at 1500mg BID with food was safe and well tolerated and a phase 1b/2a study is now being initiated in COVID-19 patients.

## Introduction

Since the emergence of coronavirus-induced disease (COVID-19) in Wuhan, China in 2019, numerous treatment candidates have been tested in late phase clinical trials. To date limited therapeutic options [1] exist, such as steroids and IL-6 receptor blockers (tocilizumab and sarilumab) as immunodulators in severe disease, and emerging signals of efficacy of antiviral monoclonal antibody therapies. Repurposing of approved drugs is in principle the fastest way to establish interventions during an urgent public health crisis. This has yielded interventions for severe COVID-19, but efforts to establish repurposed antiviral interventions have not been successful to date. Reasons for failure are multifaceted and relate to the pharmacokinetics and pharmacodynamics of the candidate antiviral drugs. In terms of pharmacokinetics, most putative repurposing drugs that have been studied preclinically are not expected to reach systemic antiviral concentrations at the approved dose and schedule [2]. Redundancy of the expected mechanism of action in vivo has also been reported, which was driven by inadequacy of the in vitro model used to demonstrate activity. For example, the primary mechanism of antiviral activity for hydroxychloroquine in Vero cells involved a process for viral entry which is secondary in vivo. As such, hydroxychloroquine activity is mitigated in animal models and in cells expressing TMPRSS2 [3, 4].

Nitazoxanide is a thiazolide FDA approved antiparasitic medicine used for the treatment of cryptosporidiosis and giardiasis [5] and also has reported activity against anaerobic bacteria, protozoa and several other viruses [6]. Rapid deacetylation of nitazoxanide in blood means that the major systemic species of the drug in vivo is tizoxanide. Tizoxanide has been shown to exhibit similar in vitro inhibitory activity to nitazoxanide for rotaviruses [7], hepatitis B and C viruses [8, 9], coronaviruses other than SARS-CoV2 and noroviruses [10, 11]. It has also demonstrated in vitro activity against influenza viruses [12, 13] and in a phase 2b/3 trial in uncomplicated influenza, nitazoxanide demonstrated a reduction in symptoms and viral shedding at a dose of 600mg twice-daily compared to placebo [14], despite these doses providing systemic concentrations that are only expected to remain above the influenza in vitro target for a fraction of the dosing interval [2]. Similarly, several recent small clinical trials have indicated some antiviral and clinical benefits of nitazoxanide but at doses that are not expected to maintain concentrations above the in vitro antiviral target for the full dosing interval [15-17]. While this is encouraging, higher doses and combinations are likely to be ultimately needed to maximise the antiviral activity and mitigate the risk of emergence of drug resistance [18, 19]. The antiviral mechanism of action of nitazoxanide against influenza involves an impact upon post-translational modification and maturation of hemagglutinin [20], and a similar mechanism involving the SARS-CoV-2 spike protein was also recently reported [21].

Other potential benefits of nitazoxanide in COVID-19 may derive from its impact upon the innate immune response that potentiates the production of type 1 interferons [13, 22] and bronchodilation of the airways through inhibition of TMEM16A ion channels [23]. Moreover, a recent study indicated that drugs which inhibit TMEM16, like nitazoxanide, block the SARS-CoV-2 spike protein-mediated syncytia formation via a mechanism independent of their antiviral activity [24]. At conventional doses of 500 mg twice daily, nitazoxanide achieves C_trough_ plasma concentrations close to the in vitro defined target concentration for SARS-CoV-2 [25], and exhibits antiviral activity in cell types that recapitulate in vivo mechanism of replication [15]. The highest nitazoxanide dose reported to date in COVID-19 clinical trials is 1000mg twice-daily, utilised in combination with another agent.

Published physiologically-based pharmacokinetic (PBPK) modelling, validated against tizoxanide pharmacokinetics after single doses ranging between 500 and 4000mg as well as twice daily doses of 0.5 and 1g, indicated doses above those already approved may provide C_trough_ above the antiviral target in the majority of patients [2]. Modelling estimated that 1400mg twice-daily or 900mg three-times daily will provide pulmonary exposures for the entire dosing interval above the reported in vitro EC_90_ for SARS-CoV-2 in over 90% of the population. Nitazoxanide has an established safety record in humans and studies showed tolerability of single oral doses up to 4000g with minimal gastrointestinal side effects. In a separate study involving 16 healthy males, doses of either 500mg twice-daily or 1000mg twice-daily for 7 days, the 500mg dose was well-tolerated with only mild adverse events not differing significantly from the placebo [26]. The 1000mg twice-daily dose was associated with an increased frequency of gastrointestinal side effects, primarily diarrhoea and abdominal discomfort, but no significant changes were noted in the safety parameters and laboratory tests.

The combination of a long-established safety track record along with its demonstrated in vitro activity against coronaviruses including SARS-CoV-2, and its availability from global generic manufacturers at extremely low cost, makes nitazoxanide an attractive therapeutic option for dose escalation and repurposing. As such it was prioritised for evaluation at higher doses within the AGILE clinical trial platform, as a potential therapeutic for mild/moderate disease.

## Methods

### Study design

This was a single-centre, open label, adaptive, phase 1 trial in healthy adult participants (NCT04746183). This was undertaken as a candidate specific trial within AGILE; a Phase Ib/IIa platform trial for evaluating new therapies for COVID-19 [www.agiletrial.net], comprising an over-arching Master Protocol [27] under which sits candidate-specific trials evaluating specific compounds. The study protocol was reviewed and approved by the UK Medicines and Healthcare products Regulatory Agency (MHRA) and West Midlands Edgbaston Research Ethics Committee. The study was coordinated by the National Institute for Health Research (NIHR) Southampton Clinical Trials Unit with participants recruited into the NIHR Royal Liverpool and Broadgreen Clinical Research Facility (UK).

### Participants

Eligible participants included healthy adult males and non-pregnant and non-lactating females between 18 and 75 years of age. Females of childbearing potential and males (who are sexually active with female partners of childbearing potential) were required to use two effective methods of contraception, one of which should be highly effective, throughout the study and for 50 days (females) and 100 days (males) thereafter. Participants were included if they were confirmed to be healthy in the absence of any clinically significant cardiovascular, respiratory, gastrointestinal, neurological, psychiatric, metabolic, endocrine, renal, hepatic, haematological or other major disorder. Specific exclusion criteria included: pregnancy or currently lactating, being in receipt of any medication, including St John’s Wort, known to chronically alter drug absorption or elimination within 30 days prior to first dose administration. In particular, owing to the high protein binding of tizoxanide; warfarin, phenytoin, amiodarone and intravenous chemotherapy were specifically prohibited within 30 days or five half-lives (whichever was longer) of first dose administration and up to the end of the study. Participants in receipt of any prescribed medication that required dose alteration or any non-prescribed systemic or topical medication, herbal remedy or vitamin/mineral supplementation within 14 days prior to the first dose administration (unless, in the opinion of the investigator, it would not interfere with study procedures or compromise safety) were specially excluded. Additionally, those with any clinically significant allergy or those that had previously received nitazoxanide or its constituent parts within 3 months of receiving first dose. All participants provided written informed consent before enrolment. The full list of inclusion and exclusion criteria can be found in supplementary Table 1. The flow of participants is outlined in Figure 1.

**Figure 1:**
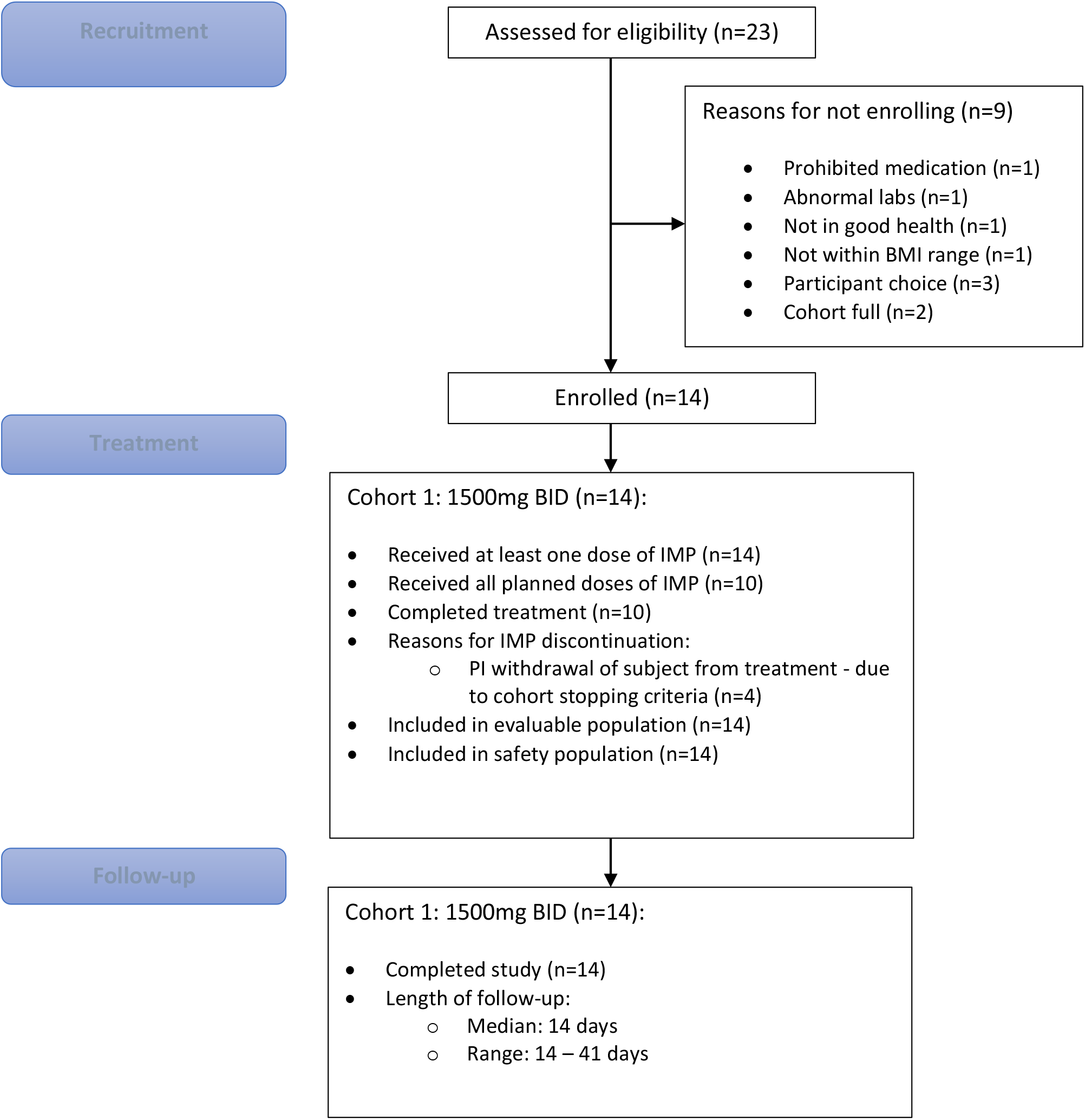
CONSORT Diagram

### Dosing

Eligible participants received 1500mg nitazoxanide orally with food, twice-daily for 7 days. Dosing with food was undertaken on the inpatient unit on day 1 and day 5; the remaining dosing was administered by the participants at home with instruction to eat within 30 minutes of dosing. In this study we had pre-defined three dosing strategies for nitazoxanide: i) 1500 mg 12 hourly (‘bd dosing’) which was predicted by our pharmacokinetic models to achieve the desired target concentrations, plus a further two regimens of ii) 1000mg 8 hourly (‘tds dosing’) and iii) 1500mg in the morning, 2000mg in the evening (‘asymmetric dosing’). A Safety Review Committee (SRC) was planned to review the initial regimen (1500 mg bd) and if deemed a failure because of toxicity or failure to achieve target plasma concentrations could recommended progression to the 1000mg tds and/or asymmetric dose cohorts. If the SRC considered a regimen as safe with an achievement of target plasma concentrations they could recommend its further investigation in a future Phase 1b/2a trial in COVID-19 patients.

### Study procedures

Viral PCR swab screening to exclude COVID-19 was performed for all participants 48-hours prior to admission to the unit. General physical examination, serum chemistry and haematology sampling were performed at screening, 48 hours prior to dosing, day 1 baseline, 72 hours and 120 hours post-dose followed by the final study visit at day 14 post-first dose. Urinalysis was performed at screening, 48 hours pre-dose, 72 hours post-dose followed by the final study visit at day 14 post-first dose. Adverse events (AEs) were collected from the time of start of the first dose and throughout the study period. For gastrointestinal tolerance the safety focus point was stool frequency and consistency assessed according to the Bristol Stool Chart (BSC).

### Outcomes

The primary outcome for this study was to assess safety, tolerability, optimum dose and dosing schedule of nitazoxanide in healthy participants through the following parameters: AEs, general physical examination, general safety assessments including ECG, vital signs, clinical laboratory analysis including urinalysis, haematology and serum chemistry. Since gastrointestinal intolerance was anticipated at the high doses of nitazoxanide used, participants were asked to fill in a Bristol Stool Chart for the duration of assessment. The primary endpoint for this trial was safety and tolerability defined by AE frequency and severity, of nitazoxanide in healthy volunteers assessed over 10 days.

Unacceptable safety and tolerability parameters were defined as:

- A treatment emergent serious adverse event (SAE) in one subject that was considered by the investigator to be related to the study drug
- Two or more adverse events, the clinical severity of which was graded by the investigator as 3 (severe), 4 (life threatening) or 5 (resulting in death)
- Two or more participants in a cohort required withdrawal due to elevated alanine aminotransferase (ALT) levels:
  - Asymptomatic elevation in ALT or AST to > three-times the upper limit of normal (ULN), confirmed by a repeat measurement within 48 to 72 hours, accompanied by either direct bilirubin level > or = to two-time ULN or International Normalised Ratio (INR) > or = 1.5, confirmed with repeat measurement within 48 to 72 hours.
  - Symptomatic elevation in ALT to >three-times ULN.
- Significant diarrhoea, defined as seven or more episodes, rated greater than or equal to five on the Bristol Stool Chart (BSC), in one day for two consecutive days was seen in two or more participants in a group

Additional participant withdrawal criteria included:

- Any clinically relevant signs or symptoms that, in the opinion of the investigator, warrant participant withdrawal.
- QTcB prolongation to >500 milliseconds or a rise in QTcB value of >60 milliseconds (whichever was lower), observed on triplicate ECGs, compared to the baseline mean QTcB.
- Positive urine drugs of abuse screen, alcohol breath rest or pregnancy test result.

### Pharmacokinetic analysis

Intensive plasma pharmacokinetic (PK) sampling for tizoxanide and tizoxanide glucuronide was undertaken on day 1 and day 5. Blood samples were collected pre-dose, 0.5, 1, 1.5, 2, 3, 4, 6, 8 and 12h after first dose administration. Additional C_trough_ PK samples were obtained at 12-hours post-dose on day 3 and 7. Tizoxanide and tizoxanide glucuronide concentrations in plasma were measured using a validated liquid chromatography tandem mass spectrometry (LC-MS) method. In brief, analytes were extracted from plasma by protein precipitation with 0.1% formic acid in acetonitrile, centrifuged and diluted in a reconstitution solution (5 mM ammonium formate: acetonitrile, 50:50 v/v) before injection onto the HPLC column. Deuterated (D_4_) internal standards were used and chromatographic separation was achieved using a reverse phase C_18_ column. The calibration range was linear between 50-45,000 ng/mL for both analytes. As per protocol, pharmacokinetic success in the trial was defined as median C_trough_ values above the 1.43 mg / L derived from the in vitro EC_90_ against SARS-CoV-2.

The combined parent tizoxanide and tizoxanide-glucuronide metabolite dataset was fitted with a parent-metabolite PK model using 1-compartment disposition for both tizoxanide and tizoxanide-glucuronide, with 1^st^ order absorption/appearance for tizoxanide. The model is similar to previous parent-metabolite PK models applied for example to rifapentine [28] and a schematic and rate equations are provided in supplementary figure 1. In the absence of definitive mass balance data for fraction of total parent drug clearance accounting for the formation route of the glucuronide metabolite, apparent CLmet and Vmet parameters were estimated for the metabolite. Fitting of this PK model to the observed plasma concentration data was carried out in the R programming environment (v 4.0.3) [29] making use of the Pracma library [30] and lsqnonlin function for nonlinear regression, treating the dataset from all participants as a naïve pool. When further data become available at the current dose, this PK model can be applied using a nonlinear-mixed effects population approach for characterisation of interindividual variability and potential covariate relationships with PK parameters.

In previously published work, a physiologically based pharmacokinetic (PBPK) model was validated for nitazoxanide using Simbiology (MATLAB R2019a, MathWorks Inc., Natick, MA, USA) and used to inform dose selection for the current trial. [31]. Therefore, a comparison was made between the observed pharmacokinetics in the trial and the a priori validated PBPK model prediction.

### Statistical analysis

We utilised a Bayesian adaptive design to support decision making in this phase Ia study. Details are provided in Supplement material. Briefly, there was uncertainty about the order of some of the dosing strategies, with respect to their toxicity. To enable us to relax the assumption of monotonicity of toxicity we used the Partial Ordering Continual Reassessment Method (POCRM) design proposed by Wages et al. [32] adjusted to the AGILE setting.

The POCRM utilises a simple, one-parameter logistic dose-toxicity model, which describes the relationship between dose-limiting toxicities (DLT) within 10 days of initiating treatment and treatment dose (1500mg BD, 1000mg TID, asymmetric dosing) and includes the uncertainty about orderings in the working model itself. For each possible ordering, the POCRM fits a CRM model and finds the posterior probability of each ordering being the true ordering. The model that is deemed the most likely one given the data observed is used for the escalation/de-escalation decisions. The prior distributions for the POCRM were calibrated to maximise the proportion of correct selection under the range of possible orderings and dose-toxicity scenarios within each ordering, where each dose strategy considered in the study was the optimum one.

At the end of the cohort, the Safety Review Committee (SRC) reviewed all available safety data including at least 10 days data for each participant in the cohort, including data on AEs, vital signs, ECG and clinical laboratory evaluations, as well as any emerging data from other studies. Following SRC review, recommendations could be to de-escalate, escalate, remain at the same dose, or continue to phase Ib. A dose was deemed to be unsafe if there was a ≥25% chance that treatment was associated with a >20% risk of dose-limiting toxicities at day 10. The model recommended the next dose-level according to which level is the most likely to correspond to DLT rate of 5-15%. However, the SRC made the ultimate decision whether to accept that the current dose was safe and met the target median C_trough_ value of 1.43 mg / L.

All analyses are reported according to CONSORT 2010 and ICH E9 guidelines on Statistical Principles in Clinical Trials. All enrolled participants were included in both the evaluable population and the safety population for analysis. Statistical analysis was undertaken in SAS version 9.4, STATA version 16 and R version 4.0.2. Baseline demographics and AEs are summarised using descriptive statistics. The estimated DLT rates for each dose strategy and equal-tail 95% credible intervals taken from the model of the most likely ordering in table 3. For active doses, we also present the probability that the DLT rate falls within 5-15% (a pre-determined acceptable target range for toxicity) and the probability of at least 20% toxicity (deemed as unacceptably toxic).

The sample size was flexible, based on the need for the study to adapt to accruing safety data. Simulations to assess model operating characteristics and to calibrate priors assumed three possible orderings and the same three dose strategies within each ordering, with cohorts of size twelve capped at a total of 36 participants.

## Results

Between 18^th^ February and 11^th^ May 2021, 14 healthy volunteers received at least one dose of nitazoxanide. Of 14 participants dosed, 10/14 (71.4%) completed 7 full days of dosing (14 doses). Four participants did not complete dosing as a result of a suspected QTcB prolongation occurring in one participant which led to suspension of dosing in the entire cohort. 4/14 discontinued dosing early; 2/4 discontinued after 6 full days (12/14 doses) and the remaining 2/4 discontinued after 1 full day (2/14 doses). All 14 participants were included in the safety and PK analyses. The flow of trial participants is shown in Figure 1. The demographics and clinical characteristics at baseline are summarized in Table 1.

**Table 1.** Participant demographics and characteristics

| Characteristics | Statistic | Nitazoxanide (1500mg twice-daily, n=14) |
| --- | --- | --- |
| Age | Median (range) | 25 (19-53) |
| Sex | Male/female % | 35.7%/64.3% |
| Ethnicity | White-English/Welsh/Scottish/Northern Irish/British | 9 (64.3%) |
|  | Any other White background | 3 (21.4%) |
|  | Any other Asian background | 2 (14.3%) |
| Body weight (kg) | Median (range) | 65.6 (61.2 – 96.3) |
| BMI (kg/m<sup>2</sup>) | Median (range) | 23.9 (19.3 – 30.8) |

### Safety results

Nitazoxanide was safe and well tolerated at 1500mg twice daily for 7 days with no significant adverse events (Table 2). Moderate gastrointestinal (GI) disturbance was seen in 3 participants (21.4%) with a further 8 participants (57.1%) experiencing mild GI symptoms (covering a spectrum of nausea, bloating, constipation, diarrhoea or loose stools and/or abdominal pain). Yellow discoloration of the urine and sclera was observed in 12 (85.7%) and 9 (64.3%) participants, respectively, without clinically significant elevation in bilirubin. This was self-limiting and resolved upon discontinuation of the drug. No grade 3 or 4 adverse events were documented.

**Table 2.** AEs and serious AEs

| Adverse event (all grades 1 or 2) | Nitazoxanide (1500mg BID) (n=14) |
| --- | --- |
| Urine Discoloration | 12 (85.7%) |
| Yellow Sclera | 9 (64.3%) |
| Nausea | 7 (50.0%) |
| Abdominal Pain | 6 (42.9%) |
| Diarrhoea | 5 (35.7%) |
| Loose Stools | 3 (21.4%) |
| Myalgia | 3 (21.4%) |
| Bloating | 2 (14.3%) |
| Headache | 2 (14.3%) |
| Right Corneal Irritation | 1 (7.1%) |
| Constipation | 1 (7.1%) |
| Flatulence | 1 (7.1%) |
| Fever | 1 (7.1%) |
| Flu Like Symptoms | 1 (7.1%) |
| Loss Of Appetite | 1 (7.1%) |
| Non-Cardiac Chest Pain | 1 (7.1%) |
| Fall | 1 (7.1%) |
| Umbilical Hernia | 1 (7.1%) |
| Dizziness | 1 (7.1%) |
| Lethargy | 1 (7.1%) |
| Dysuria | 1 (7.1%) |
| Irregular Menstruation | 1 (7.1%) |
| Semen Discoloration | 1 (7.1%) |
| Vaginal Discharge | 1 (7.1%) |
| Laryngeal Inflammation | 1 (7.1%) |
| Blister On Abdomen | 1 (7.1%) |
| Hypotension | 1 (7.1%) |
| Abdominal Gas (Rumbling) | 1 (7.1%) |
| Elevated creatine kinase | 1 (7.1%) |
| Heavy Sensation To Eyes | 1 (7.1%) |
| Leg Pain/Stiffness | 1 (7.1%) |
| Mouth Ulcer | 1 (7.1%) |
| Musculoskeletal Pain (Elbow) | 1 (7.1%) |
| Musculoskeletal Pain (Hip) | 1 (7.1%) |
| Other - Stomach Cramps | 1 (7.1%) |
| Rash | 1 (7.1%) |
| Reduced Appetite | 1 (7.1%) |
| Shortness Of Breath | 1 (7.1%) |
| Skin Discolouration | 1 (7.1%) |
| Stye (Right Eye) | 1 (7.1%) |
| Yellow Semen | 1 (7.1%) |
| **Serious adverse event (grade 3)** |  |
| Electrocardiogram QT Corrected Interval Prolonged | 1 (7.1%) |

**Table 3:** Estimated toxicity for Nitazoxanide up to day 10 from the POCRM model (for ordering 3)

| Dose level | Estimated DLT rate (95% Credible interval) | Target dose probability (5-15% DLT rate) (%) | Probability of being overly toxic (toxicity > 20%) (%) |
| --- | --- | --- | --- |
| 1500mg BID | 2.1% (0.0% - 14.4%) | 11.8% | 0.9% |
| 1000mg TID | 0.2% (0.0% - 2.1%) | 0.6% | 0.0% |
| Asymmetric | 9.0% (0.0% - 36.2%) | 26.6% | 15.2% |

Minor, self-resolving, post-dose elevation in creatine kinase (CK) was identified in 5/14 participants (maximum 869 U/L at D5 post-dose, suppl table). One of the participants reported mild localised myalgia without muscle tenderness at D5 (CK 508 U/L) which was recorded as an AE. All other elevations were not considered clinically significant.

In one participant, tachycardia (HR 112bpm) developed shortly after the second dose administration on day 1, with an ECG recorded 7 minutes following the second dose showing a corrected QTcB (Using Bazett’s formula) of 468msecs, an apparent increase of 73.5 milliseconds above the mean baseline (394.5msec). Treatment was discontinued in all being dosed. Following overnight admission for observation and serial ECGs, an apparent prolongation recurred at 5 hours and 44 minutes following the second dose, 460 millisecs (65.5 millisecs above baseline mean, HR 87bpm) and resolved fully thereafter. Independent cardiology review of all ECG’s with manual QT and QTcB calculations confirmed an artefactual QTcB increase due to the presence of a prominent U-wave fused with the end of the T wave, causing the ECG machine to calculate the QU interval instead of the QT interval. Following formal review by the trial Safety Review Committee and in consultation with the UK medicines regulator, the trial (which had been temporarily suspended) was reopened at the same dose level with replacement of 2 participants who discontinued dosing after only 1 full day. Additional ECGs were obtained at 14 hours post first-dose on D1 and D5 for subsequent participants, with no significant changes observed.

### Pharmacokinetics and modelling

Median C_trough_ at the end of the first dose were above the in vitro-defined target concentration (EC_90_ – 1.43 mg/L) and remained so throughout dosing. The simultaneous fitting of the parent-metabolite pharmacokinetic model to the tizoxanide and tizoxanide glucuronide datasets is illustrated in Figure 2, with parameter estimates and relative standard errors in Table 4. An acceptable description of the data was obtained with acceptable precision of estimates. Interindividual variability in plasma exposure appears relatively wide but may reflect to some extent variability in administration times for doses taken by patients at home compared to nominal dosing times. The PBPK simulated tizoxanide plasma concentrations relative to the naïve pool of data from healthy individuals in this study is shown over the 7days of dosing in Figure 3A. Figure 3B and 3C show the comparison of the observed and simulated median pharmacokinetic profiles on day 1 and day 5, respectively. The corresponding pharmacokinetic data and a numerical comparison between observed and simulated Cmax and C_trough_ is provided as Suppl. Table 4. The trial confirmed prior PBPK predictions for first dose but with underprediction of exposures at day 5, with higher pharmacokinetic exposures and delayed Tmax observed clinically than predicted by the PBPK modelling. Since the PBPK model was validated against the clinical data for multiple BID doses of 500mg and 1g nitazoxanide [33], where no drug accumulation was observed as the days progressed, the model was not able to capture appropriately the drug accumulation observed with the 1500 mg BID regimen with an increasing C_trough_ from day 1 to day 5 and day 7. However, concentrations above the target (i.e. EC_90_ – 1.43 mg/L) were achieved on the first dose and safely maintained throughout the course.

**Table 4.** Parameter estimates for simultaneous parent-metabolite PK model fitting to tizoxanide and tizoxanide glucuronide plasma concentration data.

| | $CL_{TIZ}/F_{TIZ}$<br>(L/h) | $V_{TIZ}/F_{TIZ}$<br>(L) | KA<br>(h<sup>-1</sup>) | $CL_{MET}/(F_{TIZ} * F_{MET})$<br>(L/h) | $V_{MET}/(F_{TIZ} * F_{MET})$<br>(L/h) |
| --- | --- | --- | --- | --- | --- |
| Parameter Estimate | 3.95 | 64.99 | 0.45 | 5.88 | 12.71 |
| % RSE* | 3.64 | 8.15 | 14.20 | 3.70 | 12.67 |

**Figure 2:**
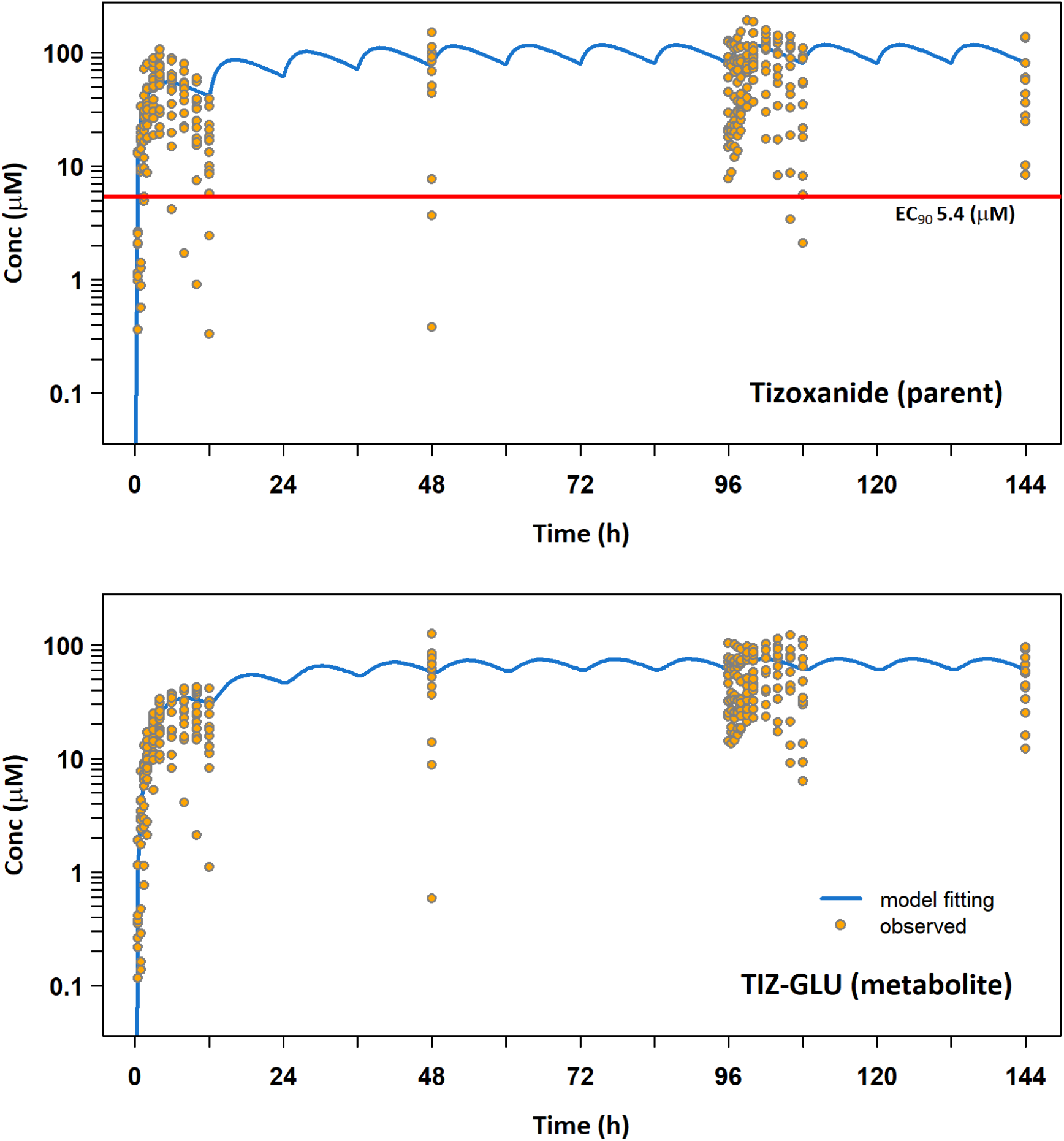
Simultaneous 1-compartment, 1^st^ order absorption parent-metabolite PK model fitting to naïve pooled tizoxanide (parent) and tizoxanide-glucuronide (metabolite) plasma concentration data. The red line represents the in vitro derived EC_90_ against SARS-CoV-2 (1.43 mg/L).

**Figure 3.**
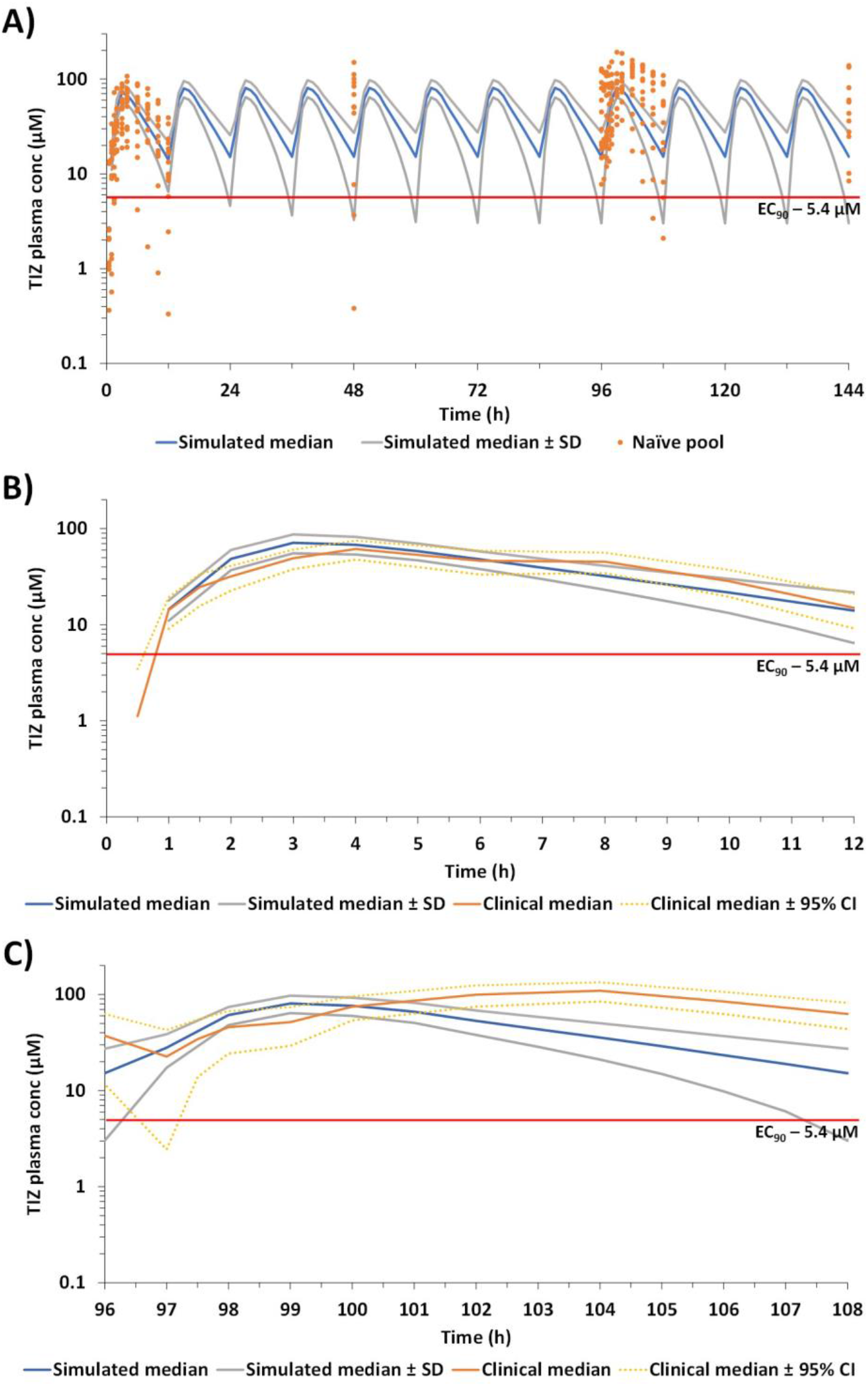
Comparison of PBPK simulated and observed tizoxanide plasma concentrations. A) comparison of median (standard deviation) simulated concentrations (blue) against the naïve pool of plasma concentration in healthy individuals. B) Comparison of observed and simulated median (95% CI) tizoxanide plasma concentrations following the first dose. C) Comparison of observed and simulated median (95% CI) tizoxanide plasma concentrations on day 5. The red line represents the in vitro derived EC_90_ against SARS-CoV-2 (1.43 mg/L; 5.4 micromolar).

## Discussion

This study establishes the safety and tolerability of high-dose nitazoxanide, with food in healthy adult participants. A nitazoxanide dose of 1500mg twice-daily for 7 days is safe and well-tolerated and the plasma concentrations attained are expected to be sufficient to achieve the in vitro-defined EC_90_ against SARS-CoV-2 in over 90% of the population. As expected, gastrointestinal effects were common (predominantly nausea, abdominal discomfort, bloating, loose stools) and at most, of moderate intensity (Grade 2 discomfort sufficient to cause interference with normal activities). Participants also reported yellow discolouration of sclera and bodily fluids in line with the Summary of Product Characteristics (SmPC) and previous reports, and these fully resolved upon discontinuation of the drug.

Minor elevation in CK, predominantly in the second half of the cohort, was noted in 5/14 participants. This was also self-resolving and not associated with muscle tenderness on physical examination. We believe increased self-reported physical activity of participants, coinciding with COVID-19 lockdown restrictions easing, may explain the mild CK elevations observed but ongoing monitoring of CK will be undertaken as part of phase Ib in COVID-19 patients.

After review, the Safety Review Committee and the AGILE Independent Data Monitoring and Ethics Committee agreed that these adverse events were minor. The POCRM model suggested an escalation to the asymmetric regimen was safe. However, the preferred starting regimen of 1500mg BD was on course to yield optimal exposure and was recommended to progress to evaluation in patients with COVID-19. An AGILE phase 1b study is now being initiated in South Africa for confirmatory PK analysis and tolerability in COVID-19 patients, with seamless transition into phase 2a.

Selection of the target plasma concentration was based upon an in vitro derived EC_90_ value generated in Vero cells using nitazoxanide and not tizoxanide [34]. However, further in vitro experimentation conducted internationally since this original report has demonstrated similar activity in lung epithelial cell models as well as activity of tizoxanide comparable to nitazoxanide itself [35-37]. Furthermore, the antiviral activity has been confirmed to involve inhibition of maturation of the SARS-CoV-2 spike protein [21], which is similar to the mechanism of antiviral activity for influenza [38]. More recently, a separate and distinct mechanism involving blocking of spike-mediated syncytia formation via interactions with TMEM16 has been reported, which may moderate disease severity in parallel to the antiviral effect [24]. Target concentrations for this secondary mechanism are yet to be clarified but warrant further investigation.

Whilst defining the optimal dose and duration of repurposed therapeutics is fundamental to later trial success, it cannot be assumed that effective doses and exposures directly translate into patients with COVID-19, particularly those with altered pathophysiology due to severe illness. This is a limitation of all first-in-human healthy volunteer studies, within which it is always difficult to generate pharmacodynamic data. The median age and lack of medical comorbidities in this healthy volunteer cohort also differs from the main target population for early use of antivirals, although their use in post-exposure prophylaxis and to reduce isolation requirements in low-risk groups is also being considered.

There is an urgent unmet need for safe and effective antiviral therapeutics in early-stage mild/moderate COVID-19. These are aimed at preventing progression of disease to hospitalisation and death, and possibly also reducing viral transmission in community settings. Re-purposing and dose escalation of nitazoxanide for COVID-19 is supported by in-vitro data, PBPK modelling and now robust safety and pharmacokinetic data at the 1500mg BD dose. This dose will provide the maximum potential to demonstrate antiviral activity of nitazoxanide in subsequent trials to provide a definitive outcome on the utility of this drug in COVID-19.

### Study Highlights

#### What is the current knowledge on the topic?

Nitazoxanide is an anti-parasitic medication licensed by the FDA at standard dosing (500mg BD) with an established safety profile. Antiviral activity has been demonstrated for numerous viruses with in vitro data demonstrating activity against SARS-CoV-2. No steady-state pharmacokinetic data are available at higher doses or in COVID-19 but PBPK modelling has indicated a 1500mg BD regimen will achieve required SARS-CoV-2 plasma EC_90_ concentrations across the dosing period.

#### What question did this study address?

Is high-dose nitazoxanide safe and well-tolerated in healthy individuals and can it achieve and maintain plasma antiviral concentrations predicted to be sufficient to prevent maturation of the SARS-CoV-2 spike protein and therefore drive antiviral efficacy?

#### What does this study add to our knowledge?

Plasma concentrations of tizoxanide, the major circulating form of nitazoxanide, are sufficient to maintain the in vitro derived EC_90_ and can be safely achieved in healthy individuals. The 1500mg BD dose is well tolerated, with mild gastrointestinal side effects in healthy volunteers.

#### How might this change clinical pharmacology or translational science?

This phase 1a study precedes a seamless Phase 1b/2a evaluation of high dose nitazoxanide in mild/moderate COVID-19 within the AGILE platform. It has provided key information on the pharmacokinetic profile and tolerability at higher doses that supports its evaluation in COVID-19 patients and potential use as an antiviral in other diseases. These doses will give the maximal opportunity to achieve antiviral concentrations for SARS-CoV-2 but the utility of nitazoxanide for COVID-19 can only be determined in subsequent trials in patients.

## Supporting information

Supplemental Bayesian design

## Data Availability

The data that support the findings of this study are available on request from the corresponding author. The data are not publicly available due to restrictions containing information that could compromise the privacy of research participants

## Acknowledgements

Sara Yeats, Emma Tilt, Emma Wrixon, Andrea Corkhill and Catherine Simpson

## Author contributions

Study design: SK, GG, TJ, PM, DL, MJ, RJF, GS, SE, TF, AO, LW

Data analysis and interpretation: SK, GG, TJ, SE, GS, KT, PM, HP, ML

Clinical conduct: LW

Clinical assessment and data collection: LW, RL, RC, TR, TF, RJF, MF

Pharmacovigilance: KF

Digital data collection and data management: RW, LJ

Bioanalysis: CH, SK, Laura Else, Sujan Dilly-Penchala, Alieu Amara

Study management and execution: HR, CW, KM, IE

Study monitoring: PM

PBPK modelling: AO, RR, HP

Primary manuscript writing: LW, TF, SK, AO, GS

All authors contributed to the final version of the manuscript.

