## Supplemental Bayesian design for "An open label, adaptive, phase 1 trial of high-dose oral nitazoxanide in healthy volunteers: an antiviral candidate for SARS-CoV-2"

Suppl. Section 1 Description of Bayesian adaptive design to support decision making

**Setting**

Let us consider a dose-schedule finding trial of the Nitazoxanide administered using different dosages and different schedules. We will refer to the combination of dosages/schedule as a regimen. There are 3 regimens to be studied in the trial which are given in Table 1.

| Regimen | Notation | Morning | Afternoon | Evening | Total Daily |
|---|---|---|---|---|---|
| BID | (1) | 1500mg | - | 1500mg | 3000mg |
| TID | (2) | 1000mg | 1000mg | 1000mg | 3000mg |
| Asymmetric | (3) | 1500mg | - | 2000mg | 3500mg |

Table 1: Treatment regimens under evaluation

The primary objective of the trial is to find the regiment that corresponds to a dose-limiting toxicity (DLT) risk of $\gamma = 10\%$. There are up to 36 patients to be enrolled into the trial in cohorts of $c = 12$ patients.

It is known prior to the trial that the toxicity of the dose/schedule increases with the dose of the drug, and with more frequent schedule (comparing the same dosage). However, the order of the dose-schedules that correspond to increase (decrease) in a dose and less (more) frequent dosing is unknown. Specifically, this implies that it is known prior to the trial that:

- BID is less toxic than Asymmetric (the same schedule but higher doses)

but unknown whether:

- TID is more or less toxic than BID;
- TID is more or less toxic than Asymmetric;

The grid of dose-schedules can be presented as in Table 2. In Table 2, a move along the rows corresponds to a change of schedule, a horizontal move corresponds to a change in the dose. Each of the `moves' from the “left to right" and from the “bottom to top" correspond to an increase in the toxicity risk.

| | | |
|---|---|---|
| TID (2) | - | - |
| - | BID (1) | Asymmetric (3) |

Table 2: Dose-Schedule grid

**Design – Model**

Given that there is uncertainty about the order of the regimens (with respect to their toxicity) concerning at least some of the regimens, a conventional single-agent model-based design is not applicable as they are based on the assumption that the doses (regimens) can be ordered with respect to increasing toxicity prior to the trial. To relax this assumption of monotonicity, we propose to employ the Partial Ordering CRM (POCRM) design proposed by Wages *et al* adjusted to the AGILE setting.

**POCRM Design – Definition**

The POCRM utilises a simple (one-parameter, working) dose-toxicity model and includes the un-certainty about monotonicity of orderings in the working model itself. Formally, assume that there are S feasible orderings of the combination/schedules. Let $\pi_{is}$, $i$ = 1, . . . ,I, s = 1, . . . ,S be the standardised combination/schedule level at level i under the ordering m, and let $p_i$ be the corresponding probability. Then, the combination/schedule-toxicity model takes the form

$$p_{is} = \pi_{is}^{\exp(\alpha_s)} \qquad (1)$$

where $\alpha_s$ is the (scalar) model parameter under the ordering s having the normal distribution $\alpha_s \sim N(0, \sigma^2)$. The working models $\pi_{is}$ are constructed from the standardised values (also known as a skeleton in the CRM) by re-ordering them according to the order s.

For each ordering s, The POCRM fits a model (1) and finds the posterior distribution of $\alpha_s$ under the ordering s. Then, given some prior distribution of how likely each of the orders are, qs, the posterior probability of each ordering being the true ordering is updated, and the ordering corresponds to the maximum, $s^*$, is found. The next cohort is allocated using model (1) for s = $s^*$.

**POCRM Design for AGILE CST-3 Phase Ia**

The POCRM design requires specification of all complete feasible orderings prior to the trial, where "complete" refers to the fact that all regimens should be contained in this ordering, and "feasible" refers to the fact that the known monotonic assumption(s) should be satisfied within these complete orderings. In the considered setting with 3 regimens and the assumption that BID is less toxic than Asymetric, there are 3 complete feasible orderings. To present the orderings for the regimens, we will use the index notation in Table 2. The orderings and the assumption that they imply are given below:

1. (1) → (2) → (3): the toxicity is primarily is driven by the total dosage;
2. (1) → (3) → (2): the toxicity is driven by the schedule;
3. (2) → (2) → (3): the toxicity is drive by the dosages of each administration

These 3 orderings will be used in the POCRM. Specifically, the single-agent CRM model will be fitted for each and then the model that is deemed the most likely one given the data observed will be used for the escalation/de-escalation decisions.

**Design - Escalation/De-escalation Rules**

The model above will be used to drive the escalation/de-escalation subject to the restrictions. The escalation restrictions are

- Escalate to safe regimens only. The regimen i is estimated to be safe if under the selected (the most likely) ordering $s^*$

$$\mathbb{P}\,(p_{is^*} > 0.20) < 25\% \qquad (2)$$

where 25% is a probability constant controlling overdosing (found by calibration to safeguard the patients - details below), and 0.20 is the upper toxicity limit such that and the regimens above 0.20 are deemed unsafe. If all regimens are unsafe, the trial is stopped earlier for safety.

- No regimens skipping is allowed (subject to the identified most likely ordering (Mozgunov and Jaki, 2019)).

The proposed design takes the following form:

1. The first cohort is allocated to BID (1);
2. After the DLT outcomes are evaluated, the probability of each ordering being the correct one are updated. The most likely ordering $m^*$is chosen.
3. The set of safe regimens satisfying the restrictions above are found.
4. Among them, the schedule i corresponding to the minimum of
   $(\hat{p}_{im^*} - \gamma)^2$ (3)
   is assigned to the next cohort of patients, where $\hat{p}_{im^*}$ estimated toxicity at schedule i under the most likely ordering $m^*$.
5. Steps 2-4 are repeated until the maximum number of patients is reached or the trial is stopped for safety (all schedules are unsafe).

**Design Parameters**

The POCRM design requires several design parameters to be specified:

- Prior variance of the model parameter, $\sigma^2$
- Skeleton values $\pi_1$, $\pi_2$, $\pi_3$
- Overdose control constant, $C_{overdose}$;
- Prior probabilities of each ordering being true, $q_1$, $q_2$, $q_3$, $\sum_i q_i = 1$.

The first three items above will be selected via a calibration procedure such that these design parameters result in high accuracy across many different scenarios. The last item, however, can be pre-specified given the interpretation/likelihood behind each of the ordering. The following rationale was used to select the ordering.

With conversations with clinicians, it was elicited that ordering 3 is deemed to be the most likely one a-priori, and ordering 2 being the least likely one. Originally, the following vector of the prior probabilities of each ordering were proposed: (0.35,0.15,0.50). Running the simulations for this prior, however, revealed that a low allocated prior probability to ordering 2 resulted in not selecting the correct regimen under scenarios when this ordering is the true one. A slight tuning to (0.30,0.20,0.50) was done to reflect the same prior clinicians' beliefs regarding the order of the likelihood of each ordering but also to resolve poor operating characteristics under some scenarios.

**Numerical Evaluation**

Below, we provide an extensive simulation study of the design proposed. As before, the maximum sample size of N = 36 and the cohort size is c = 12. The target probability is $\gamma$ = 0.10. We evaluate the proposed design in terms of the probability of the correct regimen selections.

**Scenarios**

A comprehensive simulation study should cover a wide range of regimen scenarios to ensure that the design results in high probability of accurate decision regardless of the toxicity setting. A set of 9 regimen-toxicity scenarios are given in Table 3. We denote a scenario by "Scenario xx-yy" where xx is the number of safe regimens in the scenario, and yy is the ordering which is the true one for this scenario. For example, Scenario 2-3 is the scenario with two safe regimens and the ordering 3 being the correct one. We have also included an unsafe scenario to ensure that the design can stop the trial earlier if all regimens are unsafe.

| Scenario | BID (1) | TID (2) | Asymmetric (3) |
|---|---|---|---|
| Scenario 1-1 | 0.10 | 0.25 | 0.40 |
| Scenario 2-1 | 0.01 | 0.10 | 0.25 |
| Scenario 3-1 | 0.01 | 0.02 | 0.10 |
| Scenario 1-2 | 0.10 | 0.40 | 0.25 |
| Scenario 2-2 | 0.01 | 0.25 | 0.10 |
| Scenario 3-2 | 0.01 | 0.10 | 0.02 |
| Scenario 1-3 | 0.25 | 0.10 | 0.40 |
| Scenario 2-3 | 0.10 | 0.02 | 0.25 |
| Scenario 3-3 | 0.02 | 0.01 | 0.10 |
| Unsafe Scenario | 0.35 | 0.40 | 0.45 |

Table 3: Regimen-Toxicity Scenarios

This selection of the scenario will ensure that the design can select the correct regimen regardless of the regimen location on the grid and regardless of which of the 3 considered orderings is the correct one.

**Prior Calibration**

For the design parameter discussed above, we choose those values that result in good operating characteristics under many different scenarios. The calibration over the following grid of values is performed

- **Skeleton.** The skeleton requires specification of 3 values: $\pi_1$, $\pi_2$, $\pi_3$ for 3 regimens. Importantly, the regimen (1) being the starting one and the ordering 3 being the most likely a-priori implies that the dose escalation starts in the "middle" of the regimen-toxicity relationship, so one can de-escalate (if the starting regimen is too toxic) and escalate (if the starting regimen is underdosing). This information is used to choose the value of skeleton that will be used for the starting combination (1) at the start of the trial, i.e. for the regimen in the "middle": $\pi_2$= 0:10. The values below and above this one are calibrated over the grids $\pi_1$:{0.005; 0.01; 0.02; 0.03; 0.04; 0.05} and $\pi_3$:{0:15; 0:20; 0:25; 0:30; 0:35}

- **Prior Variance** of $\alpha_m$ with the following values tried: {0:43; 0:70; 1:00; 1:34; 1:70}.

The calibration was performed over 3 scenarios generated under the ordering 1 and without applying a safety constraint. As all 3 orderings are included in the design specification, it is expected that a similar performance will be observed under different true ordering. For each combination of the values of prior parameters and under each scenario, 1000 simulations were used. The parameters resulting in the highest geometric mean of the proportion of correct selection across selection was chosen for further evaluation.

Following this procedure, the skeleton value of (0.01,0.10,0.30) and prior variance 1:34 were found to yield the highest mean proportion of correct selections. The performance of the design with these parameters is given below.

Finally, the overdosing probability $C_{overdose}$ was selected via calibration under the Unsafe Scenario. Decreasing values (starting from 0.95 with step 0.05) of $C_{overdose}$ were tried until the proportion of terminations under the Unsafe Scenario exceeded 90%. The selected value is $C_{overdose}$ = 0:25 implying that if the probability that the risk of toxicity is above 0.20 is above 25% then the regimen is deemed unsafe.

**Results**

The proportion of each regimen selections using 4000 simulations are given in Table 4.
The proportion of correct regiment selection (PCS) varies between 52% under Scenario 3-2
and 71% under Scenarios 1-3. Note that under 6 out of 9 scenarios, the PCS is above 60% and the most consistent and accurate performance corresponds to scenarios -3 that have the highest a-priori probability of being the correct one. Under the non-monotonic scenarios under which one the lower regimens are more toxic, Scenario 2-2 and Scenario 1-3, the PCS is 56% and 71% implying that the design can find the target regimen under non-monotonic schedule-toxicity relationships. Finally, the proportion of trial terminations is 93% under the highly unsafe scenario.

| | **BID (1)** | **TID (2)** | **Asymmetric (3)** | **Stop** |
|---|---|---|---|---|
| Scenario 1-1 | | | | |
| Toxicity<br>Selection | 0.10<br>**64%** | 0.25<br>18% | 0.40<br>6% | |
| Scenario 2-1 | | | | |
| Toxicity<br>Selection | 0.01<br>30% | 0.10<br>**53%** | 0.25<br>17% | |
| Scenario 3-1 | | | | |
| Toxicity<br>Selection | 0.01<br>19% | 0.02<br>19% | 0.10<br>**62%** | |
| Scenario 1-2 | | | | |

| | | | | |
|---|---|---|---|---|
| Toxicity<br><br>Selection | 0.10<br><br>**66%** | 0.40<br><br>6% | 0.25<br><br>6% | |
| Scenario 2-2 | | | | |
| Toxicity<br><br>Selection | 0.01<br><br>19% | 0.25<br><br>25% | 0.10<br><br>**56%** | |
| Scenario 3-2 | | | | |
| Toxicity<br><br>Selection | 0.01<br><br>12% | 0.10<br><br>**52%** | 0.02<br><br>37% | |
| Scenario 1-3 | | | | |
| Toxicity<br><br>Selection | 0.25<br><br>9% | 0.10<br><br>**71%** | 0.40<br><br>0% | |
| Scenario 2-3 | | | | |
| Toxicity<br><br>Selection | 0.10<br><br>**65%** | 0.02<br><br>26% | 0.25<br><br>9% | |
| Scenario 3-3 | | | | |
| Toxicity<br><br>Selection | 0.02<br><br>29% | 0.01<br><br>9% | 0.10<br><br>**62%** | |
| Unsafe Scenario | | | | |
| Toxicity<br><br>Selection | 0.35<br><br>4% | 0.40<br><br>3% | 0.45<br><br>0% | <br><br>**93%** |

Table 4: Proportion of Each Regiment Selection under 10 considered scenarios. Results are based on 4000 simulations.

**Individual Trial Behaviour**

Below, the example of the output is presented together with several cases on how the escalation will be guided by the model for the first cohort depending on the number of toxicities in the first 12 patients.

**0 DLTs out of 12**

> n
[1] 12 0 0
> y
[1] 0 0 0
### Estimated.Toxicity
[1] 0.00 0.00 0.04

### All orderings and the corresponding posterior probability
[1,] 1 2 3 36.2
[2,] 1 3 2 24.1
[3,] 2 1 3 39.7

### Overdosing probability
[1] 0.9 0.0 15.2

### Probability for the DLT rate be in (5%,15%)
[1] 11.8 0.6 26.6

### Recommended Combination
[1] 3 (escalate)

**1 DLTs out of 12**

> n
[1] 12 0 0
> y
[1] 1 0 0
### Estimated.Toxicity
[1] 0.09 0.01 0.28

### All orderings and the corresponding posterior probability
[1,] 1 2 3 28.1
[2,] 1 3 2 18.7
[3,] 2 1 3 53.2

### Overdosing probability
[1] 12.8 0.2 73.0

### Probability for the DLT rate be in (5%,15%)
[1] 46.2 8.7 13.4

### Recommended Combination
[1] 1 (stay)

**2 DLTs out of 12**

> n
[1] 12 0 0
> y
[1] 2 0 0
### Estimated.Toxicity
0.17 0.03 0.39

### All orderings and the corresponding posterior probability
[1,] 1 2 3 25.6
[2,] 1 3 2 17.1
[3,] 2 1 3 57.3

### Overdosing probability
[1] 37.4 1.5 94.6

### Probability for the DLT rate be in (5%,15%)
37.6 26.1 1.7

### Recommended Combination
[1] 2 (de-escalate)

**Example of a Figure after 1 DLT out of 12**

The example of figure presenting the mean toxicity estimates and corresponding 95% credible intervals are given in Figure 1. Note that the dots are not linked to the "dose-toxicity" relationship to avoid confusion of having a non-monotonic curve.

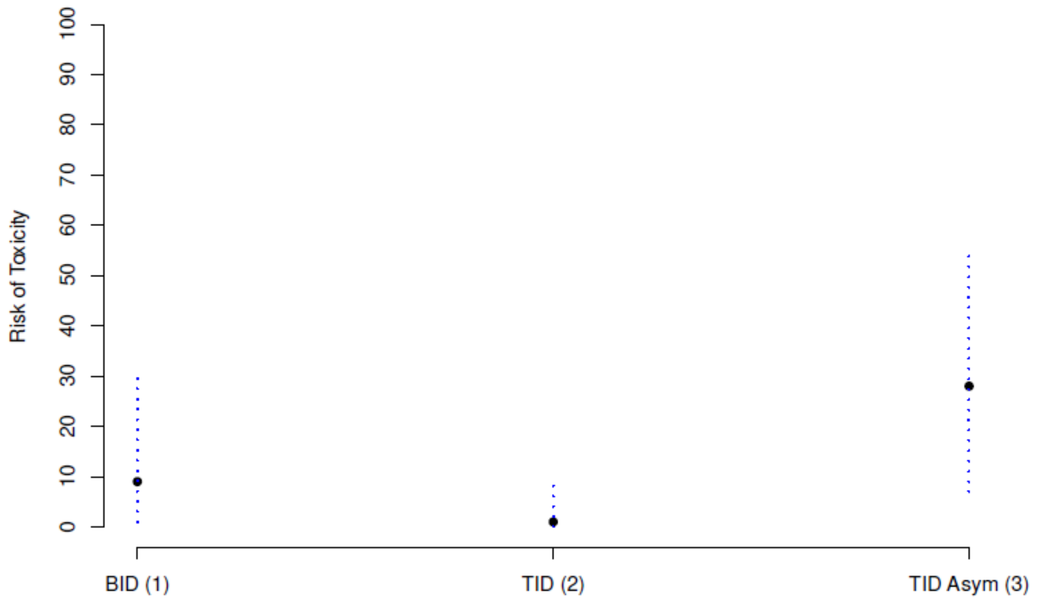
