## Supplemental Table 1 for "An open label, adaptive, phase 1 trial of high-dose oral nitazoxanide in healthy volunteers: an antiviral candidate for SARS-CoV-2"

Table S1. Full list of Inclusion and Exclusion criteria

| Inclusion criteria |  |
| --- | --- |
| 1) | Healthy Adults (≥18 years and <75 years of age). |
| 2) | Ability to provide informed consent signed by study participant. |
| 3) | Women of childbearing potential (WOCBP) and male participants who are sexually active with WOCBP must agree to use two effective methods of contraception, one of which should be highly effective and refrain from donating ova or sperm. Highly effective methods of contraception include:   - post-menopausal for 12 consecutive months - Combined Oral (estrogen and progestogen containing) hormonal contraception associated with inhibition of ovulation - Intravaginal - Transdermal - Progestogen-only hormonal contraception associated with inhibition of ovulation: a. Oral : b. Injectable : c. Implantable - Intrauterine device (IUD) - Intrauterine hormone-releasing system (IUS) - Bilateral tubal occlusion - Vasectomised partner - Sexual abstinence - Exclusive same sex relationship   The duration of contraception will be: for women, from the first administration of trial treatment, throughout trial and up to 50 days after the final dose (50 days after Day 7) and; for men with female partners of child bearing potential, from the first administration until 100 days after the final dose (100 days after Day 7) |
| 4) | In good health as defined by medical history (including confirmation from GP), physical examination, vital signs assessment, 12 lead ECG and clinical laboratory evaluation |
| 5) | BMI range 18.0-32.0 kg/m.2 |
| 6) | Written informed consent |
| Exclusion criteria |  |
| 1) | Female subjects who are pregnant or currently lactating |
| 2) | Donated blood in the 3 months prior to screening, plasma in the 7 days prior to screening, platelets in the 6 weeks prior to screening |
| 3) | Consume more than 28 units of alcohol per week if male, or 21 units of alcohol per week if female or any significant history of alcohol / substance misuse as determined by the investigator |
| 4) | Unwilling to abstain from vigorous exercise for 48 hours prior to any study visit |
| 5) | Unwilling to abstain from alcohol for 48 hours prior to any study visit |
| 6) | Received any medication, including St John’s Wort, known to chronically alter drug absorption or elimination within 30 days prior to first dose administration unless, in the opinion of the investigator, it will not interfere with study procedures or compromise safety |
| 7) | Received any medication that has required dose adjustment (with the exception of the OCP or paracetamol) within 14 days prior to the first dose administration. |
| 8) | Received any non-prescribed systemic or topical medication, herbal remedy or vitamin / mineral supplementation within 14 days prior to the first dose administration (with the exception of paracetamol) unless, in the opinion of the investigator, it will not interfere with study procedures or compromise safety |
| 9) | Subjects who have any abnormality of vital signs prior to the first dose administration that, in the opinion of the investigator, would increase the risk of participating in the study |
| 10) | Subjects who have any clinically significant abnormal physical examination finding |
| 11) | Subjects who have any clinically significant 12 lead ECG abnormality that, in the opinion of the investigator, would increase the risk of participating in the study. This includes QTcB values >450ms (male) or >470ms (female), confirmed on triplicate ECG recordings from which the mean value will be derived. |
| 12) | Subjects who have, or have any history of, any clinically significant cardiovascular, respiratory, gastrointestinal, neurological, psychiatric, metabolic, endocrine, renal, hepatic, haematological or other major disorder as determined by the investigator. |
| 13) | Subjects who have Inflammatory bowel disease (IBD) |
| 14) | Subjects who have any clinically significant allergy or allergic condition as determined by the investigator (with the exception of non-active hay fever). |
| 15) | Subjects who have any clinically significant abnormal laboratory safety results as determined by the investigator. |
| 16) | Subjects who have hepatitis B or C or are carriers of HBsAg or are carriers of HCV Ab or are positive for HIV 1/2 antibodies. |
| 17) | Subjects who have a positive alcohol breath test or a positive urine drug screen (a repeat assessment is acceptable) |
| 18) | Subjects who are still participating in another clinical study or who have participated in a clinical study involving administration of an investigational product in the 3 months (or 5 half-lives, whichever is longer) prior to first dose administration. |
| 19) | Subjects who have previously received Nitazoxanide or its constituent parts within 3 months of receiving first dose. |
| 20) | Positive for SARS-CoV-2 (using a polymerase chain reaction test) at screening and/or within 2 days prior to dosing. |
| 21) | Subjects who, in the opinion of the investigator, should not participate in this study. |
