## Supplemental Table 2 for "An open label, adaptive, phase 1 trial of high-dose oral nitazoxanide in healthy volunteers: an antiviral candidate for SARS-CoV-2"

Table S2 Summary of doses administered

| Participant | Doses administered (mg: milligrams) | Reason for discontinuation |
| --- | --- | --- |
| 01 | Nitazoxanide 1500mg twice-daily with food 14 doses, 7 days | n/a |
| 02 | Nitazoxanide 1500mg twice-daily with food 14 doses, 7 days | n/a |
| 03 | Nitazoxanide 1500mg twice-daily with food 12 doses, 6 days | Cohort stopping criteria met for participant 06 |
| 04 | Nitazoxanide 1500mg twice-daily with food 12 doses, 6 days | Cohort stopping criteria met for participant 06 |
| 05 | Nitazoxanide 1500mg twice-daily with food 2 doses, 1 day | Cohort stopping criteria met for participant 06 |
| 06 | Nitazoxanide 1500mg twice-daily with food 2 doses, 1 day | Cohort stopping criteria met due to QTcB stopping criteria |
| 07 | Nitazoxanide 1500mg twice-daily with food 14 doses, 7 days | n/a |
| 08 | Nitazoxanide 1500mg twice-daily with food 14 doses, 7 days | n/a |
| 09 | Nitazoxanide 1500mg twice-daily with food 14 doses, 7 days | n/a |
| 010 | Nitazoxanide 1500mg twice-daily with food 14 doses, 7 days | n/a |
| 011 | Nitazoxanide 1500mg twice-daily with food 14 doses, 7 days | n/a |
| 012 | Nitazoxanide 1500mg twice-daily with food 14 doses, 7 days | n/a |
| 013 | Nitazoxanide 1500mg twice-daily with food 14 doses, 7 days | n/a |
| 014 | Nitazoxanide 1500mg twice-daily with food 14 doses, 7 days | n/a |
