## Supplemental Table 3 for "An open label, adaptive, phase 1 trial of high-dose oral nitazoxanide in healthy volunteers: an antiviral candidate for SARS-CoV-2"

Table S3 Creatine kinase values

|  | Creatine kinase (CK), normal range 40-320 Units/Litre | | | | | |  | Additional clinical comments |
| --- | --- | --- | --- | --- | --- | --- | --- | --- |
| Participant | D-2 | Day 3 | Day 4^a^ | Day 5 ^a^ | Day 7 ^a^ | Day 14 | Day 28 ^a^ |  |
| 1 | 126 | 279 |  |  |  | 137 |  |  |
| 2 | 428* | 145 |  |  |  | 431* |  | Minor elevation pre-dose, regular weight training |
| 3 | 66 | 395* |  | 200 |  | 65 |  | Accidental fall on evening prior to day 3 |
| 4 | 111 | 164 |  |  |  | 214 |  |  |
| 5 | 60 | 56 |  |  |  | 46 |  |  |
| 6 | 141 | 130 |  |  |  | 128 |  |  |
| 7 | 63 | 69 |  |  |  | 73 |  |  |
| 8 | 82 | 225 |  | 265 | 348* | 90 |  |  |
| 9 | 94 | 576* | 579* | 867* |  | 101 |  | Leg stiffness on day 3, above normal exercise reported |
| 10 | 151 | 226 |  | 556* | 112 |  |  |  |
| 11 | 159 | 117 |  |  | 289 | 197 |  |  |
| 12 | 109 | 424* |  | 508* | 597* | 151 |  | Upper thigh stiffness, denied additional exercise, AE recorded |
| 13 | 76 | 234 |  |  |  | 480* | 66 |  |
| 14 | 117 | 253 |  | 869* | 359* | 117 |  |  |
