## Supplemental Table 4 for "An open label, adaptive, phase 1 trial of high-dose oral nitazoxanide in healthy volunteers: an antiviral candidate for SARS-CoV-2"

Table S4. Observed tixoxanide pharmacokinetic parameters (Clinical median (range – 95% CI) and comparison with a priori validated PBPK simulated parameters.

| **Parameter** | **Day 1 (0-12 h)** | **C_48h_** | **Day 5 (96-108 h)** | **C_144h_** |
| --- | --- | --- | --- | --- |
| **C_max_ (µM)** | 61.5 (47.6 – 75.3) | - | 109.4 (84.8 – 134) | - |
| **C_min_ (µM)** | 14.9 (9.13 – 20.7) | 51.1 (27.7 – 74.5) | 37.3 (11.7 – 62.9) | 43.8 (20.3 – 67.3) |
| **AUC_0-12_ (µM.h)** | 569.1 | - | - | - |
| **T_1/2_ (h)** | 5.0 – 6.0 | - | 4.0 – 6.0 | - |
| **T_max_ (h)** | 4.0 | - | 6.0 | - |
| **C_min_**  **(fold change, Obs. vs. PBPK)** | 1.06 | 3.37 | 2.46 | 2.89 |
| **C_max_**  **(fold change, Obs. vs. PBPK)** | 0.86 | - | 1.35 | - |
