## Supplemental Figure 1 for "An open label, adaptive, phase 1 trial of high-dose oral nitazoxanide in healthy volunteers: an antiviral candidate for SARS-CoV-2"

Figure S1: Schematic and equations of PK model for Tizoxanide and Tixoxanide-Glucuronide plasma concentrations, fitted to observed exposure data.

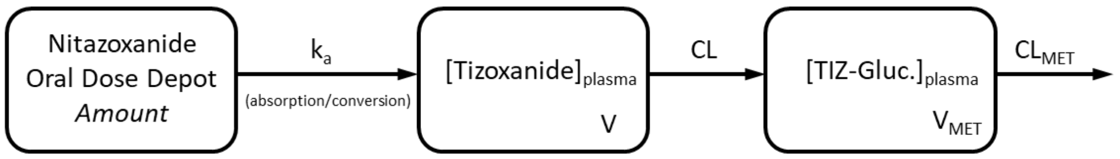


$$d(NTZ_{Depot})/dt = -\,ka * NTZ_{Depot}$$

$$d([Tizoxanide]_{plasma})/dt = +\,ka * NTZ_{Depot}/V \;-\; CL * [Tizoxanide]_{plasma}/V$$

$$d([TIZ\text{-}Gluc.]_{plasma})/dt = +\,CL * [Tizoxanide]_{plasma}/V_{MET} \;-\; CL_{MET} * [TIZ\text{-}Gluc.]_{plasma}/V_{MET}$$

[ ] denotes concentration. ***Apparent*** CL and V parameters estimated as follows, given unknown bioavailability and fraction of total CL accounted for by this metabolic route: $CL = CL/F_{TIZ}$ , $V = V/F_{TIZ}$ , $CL_{MET} = CL_{MET}/(F_{MET}*F_{TIZ})$ , $V_{MET} = V_{MET}/(F_{MET}*F_{TIZ})$
